# A capacity assessment of healthcare workers to provide second-line antiretroviral therapy in Malawi: 2016

**DOI:** 10.1101/2021.10.20.21265306

**Authors:** Wingston Ng’ambi, Yusuf Babaye, Paul Nyasulu, Andreas Jahn, Thoko Kalua, Abiba Longwe-Ngwira, Michael Odo, Ireen Namakhoma, Rose Nyirenda, Gabrielle O’Malley

## Abstract

**INTRODUCTION:** According to the estimates by the World Health Organisation, Malawi is lagging behind for the number of patients that should have been on second-line ART. In settings like Malawi the low switching may be attributable to low knowledge of clinical assessment for first-line antiretroviral treatment (ART) failure. We analyzed the knowledge level of different cadres of health workers on the management of second-line ART.

**METHODS:** This cross-sectional study assessed whether the first-line ART providers were capable to provide second-line ART in Malawi. Descriptive statistics were conducted using STATA v15.0. The passing score was 80%. Associations were assessed using Chi-Square tests with a statistical significance set at P<0.05.

**RESULTS:** Of the 754 ART providers assessed, 290 (38%) were eligible to prescribe second-line ARVs. We observed significant differences for eligibility by cadre and district (P<0.001). Furthermore, 69% of the ART providers correctly identified second-line ARVs while 9% of the providers correctly chose second-line ARVs for management of different side-effects. Both competencies varied by cadre and district (P<0.001). Of the ART providers, 9% correctly interpreted the VL results which we found to significantly vary by cadre (P<0.001) and not districts. However, 78% of ART providers were able to prescribe correct dose of second-line ART which did not significantly vary by cadre (P=0.27). Spatial analysis showed Thyolo and Chiradzulu as high performing districts.

**CONCLUSIONS:** This assessment found that registered nurses had comparable knowledge to medical doctors with at least Bachelor of Medicine and Bachelor of Surgery and clinical officers with a Diploma in Clinical Medicine in most areas but not in correctly selecting second line ARVs. To strengthen task shifting approaches, capacity building should focus on how to correctly select second line regimen and interpret viral load results. Training activities should also consider regional and district-level variability in capacity.

## BACKGROUND

Malawi is one of the few high HIV burdened countries that is on track to achieving the Joint United Nations Programme on HIV/AIDS (UNAIDS) 90-90-90 targets, with an estimated one million people living with HIV (PLHIV), 91% of whom know their status, 83% of those who know their status were on ART and 90% of those on ART were virally suppressed [1]. Antiretroviral therapy (ART) has been free for PLHIV in Malawi since 2004. As of December 2019; there were 805,232 people on ART [1]. The WHO estimated that around 5.2% of the patients receiving first-line ART should have been on second-line treatment [3]. However, as of June 2016, only 1.5% of the patients receiving ART in Malawi were receiving second-line antiretroviral drugs [2]. Similar low rates of switching patients to second-line ARV regimens are typical of most sub-Saharan African countries[4].The low switching rates are due to unavailability of viral load results [4], longer turnaround time for VL results from VL reference laboratories, and lack of proper second-line ART training for providers.

Switching to second-line ART largely depends on careful clinical assessment and access to biological measurements [4]. Misclassification of virological ART failure has frequently been reported when using WHO clinical and immunological criteria of ART failure in poor settings. As a result, Malawi adopted the use of viral load tests for confirming virological ART failure [6]. Second-line ART in Malawi has been associated with substantial mortality, morbidity and toxicity but, among survivors, virological outcomes were favourable [7] [8].

In 2016, the International Training and Education Center for Health (I-TECH) in collaboration with the Ministry of Health (MoH) and other U.S. President’s Emergency Plan for AIDS Relief (PEPFAR) implementing partners conducted a certification exam to assess knowledge of health care workers who were providing first-line ART and to ascertain their readiness to start providing second-line ART following a national refresher training. All ART service providers undergo refresher training as national HIV guidelines evolve, including updates on management of second-line ART. At the end of refresher trainings, ART providers are assessed on ability to provide second-line treatment before being certified to do so.

The national assessment of the capacity of the first-line ART providers’ readiness to offer second-line ART was done in 2016 for the first time. The assessment was done in order to increase the number of second-line ART providers in Malawi and there was no prior analysis of similar small-scale assessments that were implemented by the MoH HIV/AIDS programme implementation partners in Malawi. Therefore, in this study, we conducted analyses of the collected assessment data to evaluate the knowledge of different cadres of health workers on the management of second-line ART. We specifically assessed ART providers’ knowledge on: a) management of second-line ARV side-effects and contra-indications; b) identifying correct second-line antiretroviral (ARV) regimens; c) interpretation of HIV viral load (VL) results and adherence to HIV viral load testing protocols; and d) dosing of second-line ARV regimens.

## METHODS

### Study design

We conducted a retrospective review of cross-sectional data for the second-line ART certification examination results of first-line ART providers in Malawi in 2016. Only first-line ART providers that participated in the second-line ART prescription training and certification examinations were included in this study.

### Study setting

This was a nationwide knowledge assessment of ART providers trained to provide second-line ART in Northern, Central and Southern regions of Malawi. All first-line ART providers were invited to five-day second-line ART training workshops conducted throughout 2016. The main purpose of these training workshops was to increase the number of second-line ART providers. A total of 28 second-line ART workshop sessions with 754 participants were conducted. The workshops included participants from all the 26 districts in Malawi. At the end of each workshop, a standardized certification exam was administered to the participants. All 754 ART providers who had participated in the second-line workshops took the certification exam.

### Viral Load monitoring in Malawi

The management of patients on ART is done in accordance to both the national and international ART guidelines [10] [11] [13]. All patients that are on first-line ART are eligible for VL at 6 months and then 24 months post ART initiation date and thereafter VL was done every two years by 31 December 2016 [10]. However, if a doctor or clinician suspects ART failure, a targeted VL is requested[10]. Any viral load result of at least 1000 is suggestive of ART failure if the patient’s ART adherence has been good for the last three months over which the VL monitoring for the patient was done[12]. Therefore, such a patient gets switched to second-line ART[10].

### Second-line ART certification examination

The second-line capacity assessments were being coordinated by the Department of HIV/AIDS under the Malawi Ministry of Health. The same set of questions were provided to a team of interested first-line ART providers in 2016. This assessment was administered simultaneously across several districts of Malawi. After completion of the assessment exercise in each district, the District ART Coordinator was expected to send the marked scripts to the DHA Offices in Lilongwe. Marking of the assessment examination was done by the expert clinicians and nurses also known as ART mentors. The assessment was done as a results of ART guideline change to ensure that the first-line providers were ready to provide care to the second-line ART patients. Those who passed according to the minimum mark set by the DHA were certified by the DHA to provide second-line ART in Malawi.

### Variables

The outcomes of the study are proportions of ART providers that correctly: a) selected second-line ARV regimens, b) identified second-line ARV regimens, c) interpreted laboratory results, d) dosed second-line ARV regimens. In addition, we estimated the proportion of cadres that got at least 80% in their second-line ART certification examination. Each of the twenty-five questions had an equal weight (i.e. 4×25=100). The 80% cut-off was arrived at as a programmatic decision by the MoH. The predictors are: district of workplace, cadre (medical doctor, clinical officer, medical technician, medical assistant, registered nurse, nurse midwife technician and community nurse) and region of workplace.

### Data management

A standard, paper-based exam with twenty-five questions was administered to each participant as part of the knowledge assessment to provide second-line ART. The data were double-entered in Census and Survey Processing System (CSPro) version 7 (United States Census Bureau, Suitland, Maryland, U.S.). Double data entry was conducted by two data clerks to minimize errors due to data entry [13]. The two copies were compared and all discrepancies were resolved by referencing the hard-copy of examination paper.

The twenty-five questions were categorized into the following domains: a) identifying names of ARV regimens, b) regimen selection and identification of ARV side-effects, c) regimen dosing, and d) interpretation of viral load results and adherence to viral load testing protocols.

The data were exported from CSPro to STATA 15.0 (Stata Corp., College Station, Texas) for further data management and analysis. Within each domain, we calculated the proportion of health workers that correctly answered all the questions. The questionnaire has been provided as a supplementary file (Supplimentary_file_1.pdf).

#### Box 1: Mapping of questions from the questionnaire to each of the theme

| **Question Number** | **Theme or domain** |
| --- | --- |
| Questions included are: 1, 2, 3 and 4. We then generated a total of the correct scores. If some one had a score of four then he passed this domain. | **Correctly identified second-line ARV regimens** |
| Questions included are: 5, 7, 8, 9, 13, 19, 20 and 25. We then generated a total of the correct scores. If someone had a score of eight then he passed this domain. | **Correctly selected second-line ARV regimens for managing different patients** |
| Questions included are: 10, 11 and 12. We then generated a total of the correct scores. If someone had a score of three then he passed this domain. | **Correctly dosed different second-line ARV regimens** |
| Questions included are: 6, 14, 15, 16, 17, 18, 21, 23 and 24. We then generated a total of the correct scores. If someone had a score of nine then he passed this domain. | **Correctly interpreted viral load results** |

### Statistical analysis

Data were analysed using frequencies and percentages. We presented the results categorically with those scoring ≥80% as having passed and certified for second-line ART management; whereas within each domain we expected the ART providers to get all the questions right (i.e. pass mark of 100%). We assessed association between ART providers’ characteristics and outcomes of this assessment using chi-square test. Proportions of staff by cadre that correctly answered the questions based on the domains were spatially presented using QGIS[14]. Levels of statistical significance was set at 5%.

### Ethical considerations

Ethical approval for this assessment was obtained from the National Health Science Research Committee (Lilongwe, Malawi), University of Washington (Seattle, WA), and the Centers for Disease Control and Prevention, Center for Global Health Associate Director for Science (Atlanta, GA).

## RESULTS

### ART provider characteristics and number of certified second-line ART providers

The characteristics and percentage of ART providers certified to prescribe second-line ART are shown in Table 1. Of the 754 ART prescribers who took the certification exam, 400 (53%) were from southern, 220 (29%) were from northern and 134 (18%) were from central regions of Malawi. Further, 21 (3%) were medical doctors with at least Bachelor of Medicine and Bachelor of Surgery, 241 (32%) were clinical officers with a Diploma in Clinical Medicine with a Diploma in Clinical Medicine, 260 (34%) were nurse and midwife technicians, 147 (20%) were medical assistants and the rest (11%) were nurses of other categories (Table 1).

A total of 290 (38%) participants passed the overall second-line ART certification exams. A higher percentage of ART providers meeting certification standards were from northern region (40%) than from southern region (34%), though the difference was not statistically significant. (see Table 1). There were statistically significant differences in the proportion of ART providers passing second-line ART certification exams by cadres (P<0.001) (see Table 1). The medical doctors with at least Bachelor of Medicine and Bachelor of Surgery had the highest proportion of ART providers that passed the second-line ART certification examinations. The medical assistants had least proportion of ART providers that passed second-line ART certification examinations. Slightly over half of the clinicians passed second-line ART certification exams. Among all the ART providers; registered nurses, community nurses, clinical officers with a Diploma in Clinical Medicine and medical doctors with at least Bachelor of Medicine and Bachelor of Surgery had similar proportions of certified second-line ART providers since their 95% CI for the certified second-line ART providers overlap. However, the 95% CI for medical assistants did not overlap with the 95% CI for the certified medical doctors with at least Bachelor of Medicine and Bachelor of Surgery or clinical officers with a Diploma in Clinical Medicine.

Figure 1 shows that there were significant differences in the percentages of ART providers being certified to provide second-line ART from the 26 districts in Malawi (P<0.001). In Thyolo, Chiradzulu and Dowa districts between 61 and 76 percent of the ART providers were certified to provide second-line ART. Karonga, Kasungu, Lilongwe, Dedza and Phalombe districts had between 40 and 60 percent of the ART providers certified to provide second-line ART.

### Identification of second-line ARV regimens

Of the 754 ART providers, 518 (69%) correctly identified all the second-line ARV regimens (see Table 1). The highest percentage of ART providers correctly identifying second-line ARV regimens was from the southern region while the lowest was from the central region. The percentages of ART providers identifying the second-line ARV regimens did not statistically significantly differ by region of duty station (P=0.06). Similar proportions of medical doctors with at least Bachelor of Medicine and Bachelor of Surgery compared to clinical officers with a Diploma in Clinical Medicine, medical assistants, registered nurses and community nurses correctly identified the second-line ARV regimens. However, the 95% CI correctly identifying second-line ARV regimens for the clinical officers with a Diploma in Clinical Medicine and nurse midwife technicians do not overlap. The proportion of ART providers correctly identifying the second-line ARV regimens was highly unlikely to have occurred by chance.

Figure 2 shows that between 81 to 100% of the ART providers from Dowa, Balaka, Thyolo and Chiradzulu districts correctly identified all the second-line ARV regimens. Mchinji district had the least percentage of ART providers that identified all the second-line ARV regimens. There was very strong association between district and percentage of ART providers correctly identifying second-line ARV regimens.

### Selecting appropriate regimens for managing ARV-related side-effects

Of the 754 ART providers, 71 (9%) correctly identified regimens for managing different patient conditions related to second-line ARV regimens. We observed similar proportions of ART providers selecting appropriate second-line ARV regimens for the three regions (see Table 1). However, there were statistically significant differences among the ART providers’ cadres who correctly selected appropriate second-line ARV regimens: 17% of the clinicians (95% CI:12.13-21.91%) correctly selected appropriate second-line ARV regimens while no registered nurse selected appropriate second-line ARV regimens (P<0.001).

Figure 3 shows that there was very strong evidence that the ARV providers that selected appropriate second-line ARV regimens differed by district (P<0.001). Thyolo district had the highest percentage of ART providers that correctly selected the second-line ARV regimens for managing different patient conditions. Lilongwe, Blantyre and Mulanje districts had between 15 and 21% of the ARV providers that correctly selected the second-line ARV regimens for managing different patient conditions.

### Dosing different second-line ARV regimens

A total of 588 (78%) ART providers used correct dosing for second-line ARV regimens (see Table 1). We observed strong evidence of association between region of duty station and correct dosing of second-line ARV regimens. There was weak evidence of association between ART providers’ ability to use correct dose of the second-line ARV regimens and cadre of ART provider.

Figure 3 shows that there was strong evidence of correct dosing of various second-line ARV regimens by district (P<0.001). Salima and Rumphi districts had the least percentages of the ART providers that applied correct dose of second-line ARV regimens. Nkhata Bay, Lilongwe and Chiradzulu districts were the districts with the highest percentages of ART providers that applied correct dose of the second-line ARV regimens.

### Interpretation of viral load results

Sixty-seven (9%) of ART providers correctly interpreted the viral load results (see Table 1). There was very weak evidence of statistical significance between the proportion of ART providers correctly interpreting VL results and ART providers’ region of duty station (P=0.81). Whereas 15% of the community nurses and clinical officers with a Diploma in Clinical Medicine correctly interpreted viral load results, 14% of medical doctors with at least Bachelor of Medicine and Bachelor of Surgery correctly interpreted VL results (see Table 1) and we observed strong association between correct interpretation of viral load results and cadre of ART providers (P<0.001).

Figure 5 shows that Thyolo and Chiradzulu had between 17% and 23% of ART providers that correctly interpreted VL results as well as knowing when to perform virological testing. Kasungu and Phalombe districts had between 11 and 17% of the providers that correctly interpreted VL results and knew when to test for HIV viral load. There was weak evidence of association between district and knowledge of viral load result interpretation and testing (P=0.27).

## DISCUSSION

This is the first national study analysing the level of second-line ART knowledge among ART providers in Malawi. Overall, performance across all the domains was sub-optimal, with only 38% meeting certification requirements. Although medical doctors with at least Bachelor of Medicine and Bachelor of Surgery had the highest level of knowledge across the domains assessed, registered nurses had comparable knowledge to medical doctors with at least Bachelor of Medicine and Bachelor of Surgery and clinical officers with a Diploma in Clinical Medicine in most of the assessed areas providing potential for task shifting provision of second-line ARVs using the registered nurses. Further, there was high knowledge level among ART providers to correctly identify and use correct dosing of the second-line ARV regimens. Providers struggled with selecting appropriate regimens for managing ARV-related side-effects as well as interpreting VL results. Regimen dosing was the only variable that recorded statistical significance with region of duty station. Further, Chiradzulu and Thyolo districts had the ART providers with highest level of knowledge on most of the assessed areas.

With the overall sub-optimal performance of second-line ART certification examination, the Malawi Department of HIV and AIDS (DHA) should consider providing more in-service training on second-line ART prescription in order to have well-trained second-line ART prescribers[15]. This will then lead to reduction in the unmet need for second-line ART uptake among the patients on first-line ART[3]. Further, targeting the ART providers with more enhanced ART refresher trainings, enhanced supportive supervision and enhanced clinical mentoring may also help increase the level of knowledge on ARV medicines and management of ART patients [22]. MoH and Christian Health Association of Malawi with support from I-TECH have developed HIV modules for inclusion in pre-service training curriculum of health care workers (HCWs) in Malawi. This could serve as another platform to strengthen the competency of different cadres of HCWs in management of second-line ART.

In view of the fact that the overall level of knowledge between the clinical officers with a Diploma in Clinical Medicine, nurses and medical doctors with at least Bachelor of Medicine and Bachelor of Surgery were similar, this provides an opportunity for task shifting on initiating second-line ART to clinical officers with a Diploma in Clinical Medicine and nurses[21]. This is further supported by the high level of knowledge by these cadres on regimen identification and dosing. Similar to a study conducted in rural Malawi[15], we also found similar level of knowledge between the nurses, clinicians and medical doctors with at least Bachelor of Medicine and Bachelor of Surgery on ART patient management. It is noteworthy that nurses provide first interface in HIV care and see as much as 75% of the patients in some of the facilities of Malawi. Furthermore, the Malawi Ministry of Health has more nurses providing ART than clinicians and medical doctors with at least Bachelor of Medicine and Bachelor of Surgery. Introducing a nurse- or clinician- initiated second-line ART could be essential for scale-up of second-line ART in Malawi. However, this will require desirability of supportive supervision, mentoring, and monitoring and evaluation to strengthen implementation [15] [18].

As observed in this study, the MoH should strengthen trainings on VL monitoring since most of the ART providers scored poorly when assessed on time of collection of VL and interpretation of VL results in patient management. Without proper knowledge on this, achievement of the third 90 in the UNAIDS 90-90-90 remains elusive. ART providers should have increased knowledge on identifying which patients are failing on first-line ART and what second-line ARV regimens should be given to patients. There is also need for the DHA to provide adequate and sustainable second-line ART training to ART providers [19]. In addition to generalized ART trainings, it may be necessary to implement some specific trainings according to regional needs. For example, one of the focus areas of second-line ART trainings in the northern region of Malawi should be on regimen dosing since the least proportion of ART providers from this region correctly dosed second-line ARVs.

With the highest knowledge of ART providers from Chiradzulu and Thyolo districts, there is need for partners of DHA to learn from these districts. This highest knowledge level in these districts could be due to enhanced support from implementation partners and dynamic District Health Office leadership which could be replicated in other regions. The Malawi HIV Care and Treatment Technical Working Group could be used as a forum for the two districts to share best practice so that other districts can draw lessons from Chiradzulu and Thyolo districts on why these districts have ART providers with superior knowledge on provision of second-line ART.

One of the limitations of this study include some of the data not being collected like age of the first-line ART provider and years of service by the provider. There were also none or few numbers of ART providers to be assessed from Nkhotakota, Nasnje, Zomba and Likoma. This may have been due to differential in transport logistical arrangements of the assessment forms from these districts to Lilongwe hence having these forms arriving after data entry of data from the other districts. Finally, it took longer to have the data cleared for analysis hence we have had a longer time lag between the time when data were collected and analysis submitted as a paper for peer review. However, we still expect these findings to inform the HIV management especially of the second-line ART patients in Malawi and other similar settings.

## CONCLUSION

The findings in this study imply that it is mostly medical doctors with at least Bachelor of Medicine and Bachelor of Surgery, clinical officers with a Diploma in Clinical Medicine s and registered nurses that had similar level of knowledge to provide second-line ART. For task shifting to work well, there is need for more second-line trainings for ART providers. Most ART providers have difficulties in interpreting VL results, hence the need to specifically concentrate on improving this skill among all second-line ART providers. Geospatial analysis showed Thyolo and Chiradzulu as high performing districts. The Malawi MoH should consider the two districts to share best practice so that other districts can draw lessons from Chiradzulu and Thyolo districts on why these districts have ART providers with superior knowledge on provision of second-line ART. High HIV prevalent settings should embark on routine assessment of the level of knowledge of the ART providers in order to achieve the UNAIDS 90- 90-90 HIV strategy. There is also a need to conduct a more detailed assessment that should capture most of the variables to enable the conduct of a multivariate analysis of the assessment by adding in the assessment forms more data variables including open-ended questions that would help enhance the assessment results.

## Data Availability

Data should be accessed from the Corresponding Author

## List of abbreviations

ART: Antiretroviral Therapy
ARV: Antiretroviral Drugs
CDC: United States Centers for Disease Control and Prevention
CSPro: Census and Survey Processing System;
DHA: Department of HIV/AIDS
I-TECH: International Training and Education Center for Health
HCWs: Health Care Workers
HIV: Human Immunodeficiency Virus
MoH: Ministry of Health
PEPFAR: U.S. President’s Emergency Plan for AIDS Relief
UNAIDS: United Nations Programme on HIV/AIDS
VL: Viral Load
WHO: World Health Organisation

## Availability of data and materials

The data and do-file for this study are available online together with the article.

## Declarations

We declare that there is no conflict of interest in publishing this paper.

## ACKNOWLEDGEMENTS

The authors would like to thank William Daelo for his contribution in data cleaning for this project. The authors would also like to thank the staff in the 26 districts that participated in the second-line ART certification examination. Funding for this project was provided to I-TECH-Malawi by the United States Centers for Disease Control and Prevention (CDC). During the study period, WN, PN, MO, AJ, ALN, SG, IN, GO and YB worked for a CDC funded project. The funder had no role in the study design, data collection and analysis, decision to publish, or the presentation of the manuscript.

## Notes

### Competing Interest Statement

The authors have declared no competing interest.

