## Supplementary material for "A capacity assessment of healthcare workers to provide second-line antiretroviral therapy in Malawi: 2016": Tables and Figures

**Figure 1: Percentage of certified second-line ART prescribers in Malawi in 2016 (n=743)**

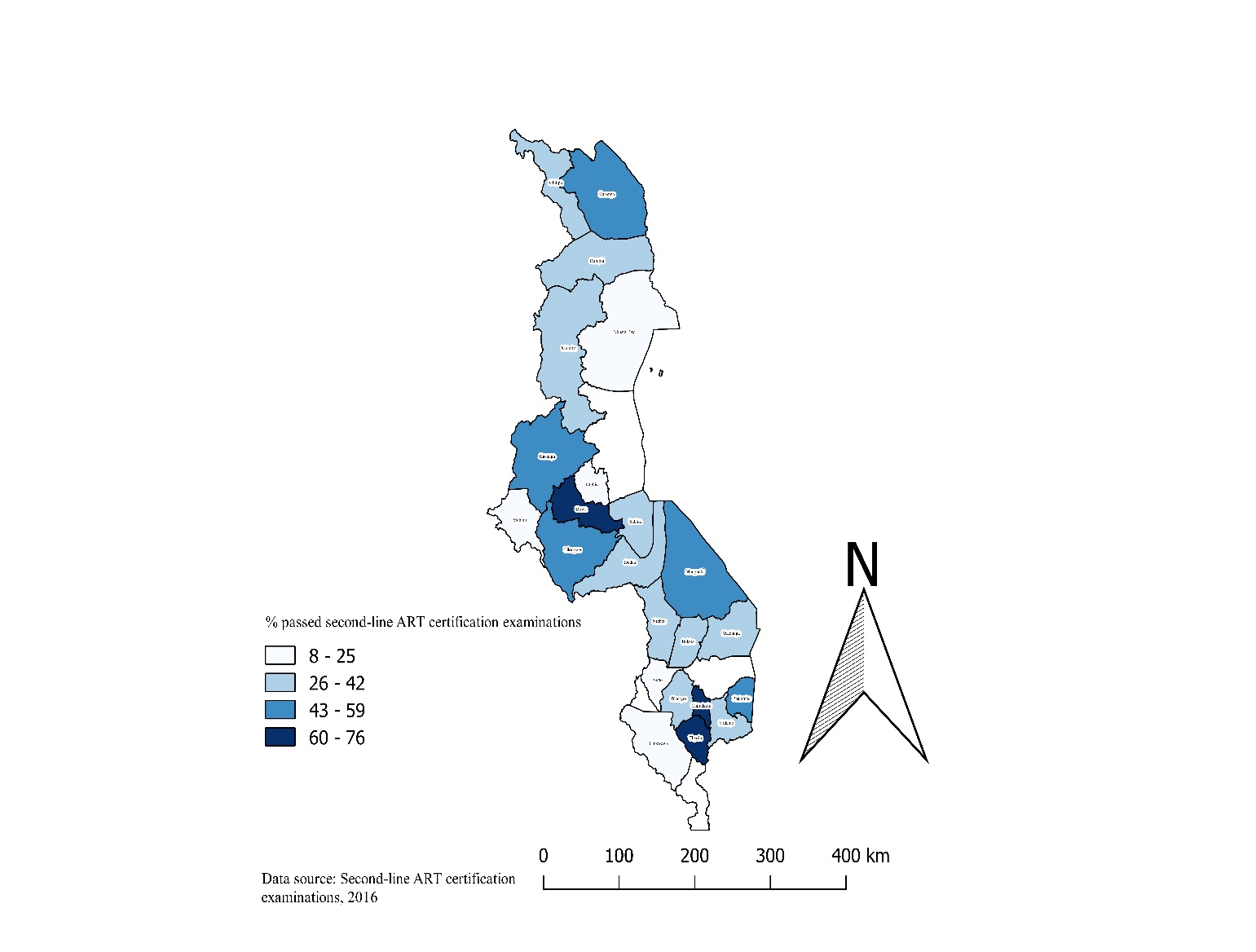

ART=antiretroviral therapy. We excluded the following districts due to small numbers: Zomba (n=7), Nsanje (n=3), Nkota-kota (n=1)

**Figure 2: Percentage of ART prescribers that identified all the second-line ARV regimens in 2016 in Malawi (n=743)**

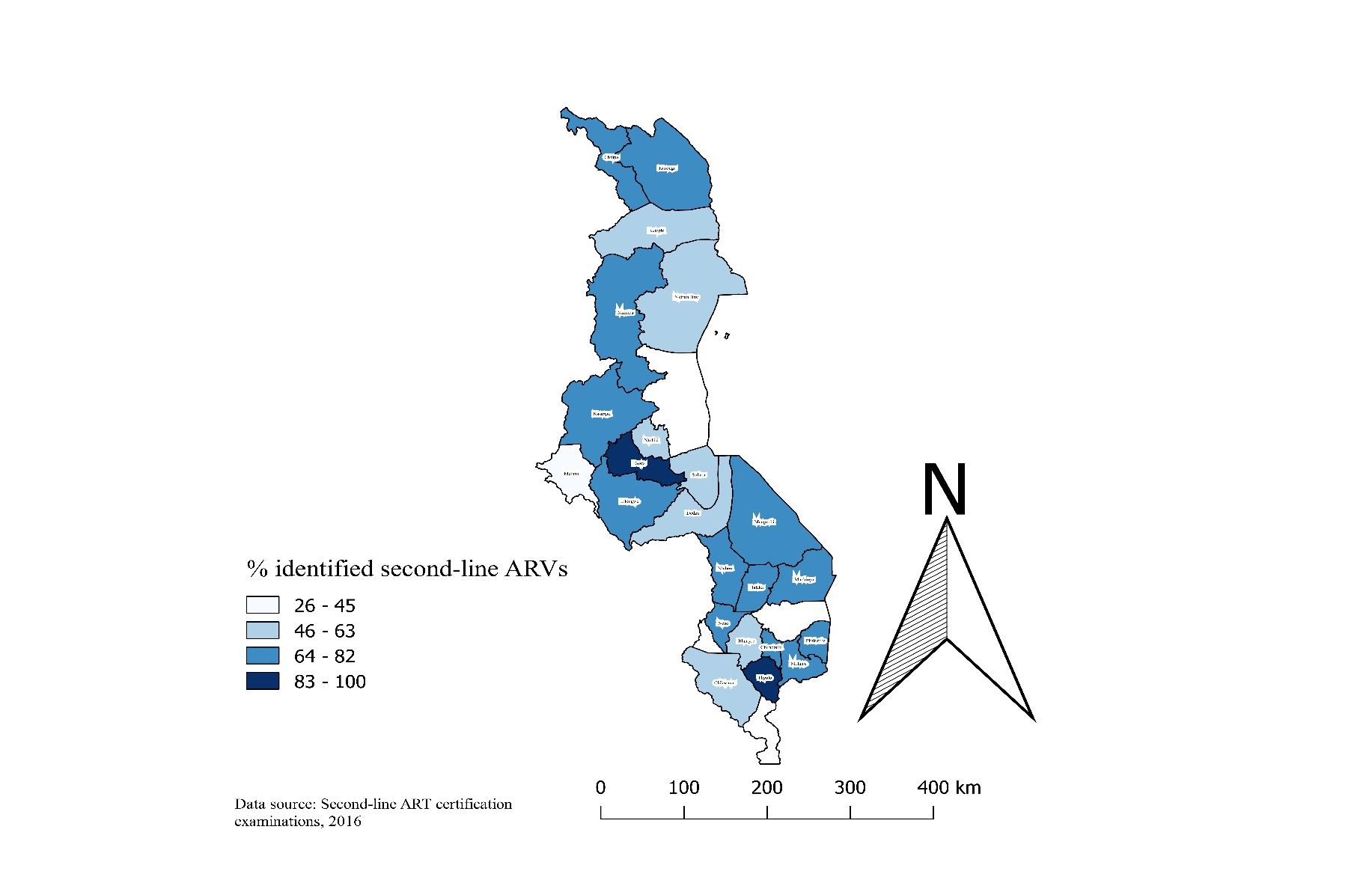

ARVs=antiretroviral drugs. We excluded the following districts due to small numbers: Zomba (n=7), Nsanje (n=3), Nkota-kota (n=1)

**Figure 3: Percentage of ART prescribers that selected appropriate second-line ARV regimens for managing different patient conditions in Malawi in 2016 (n=743)**

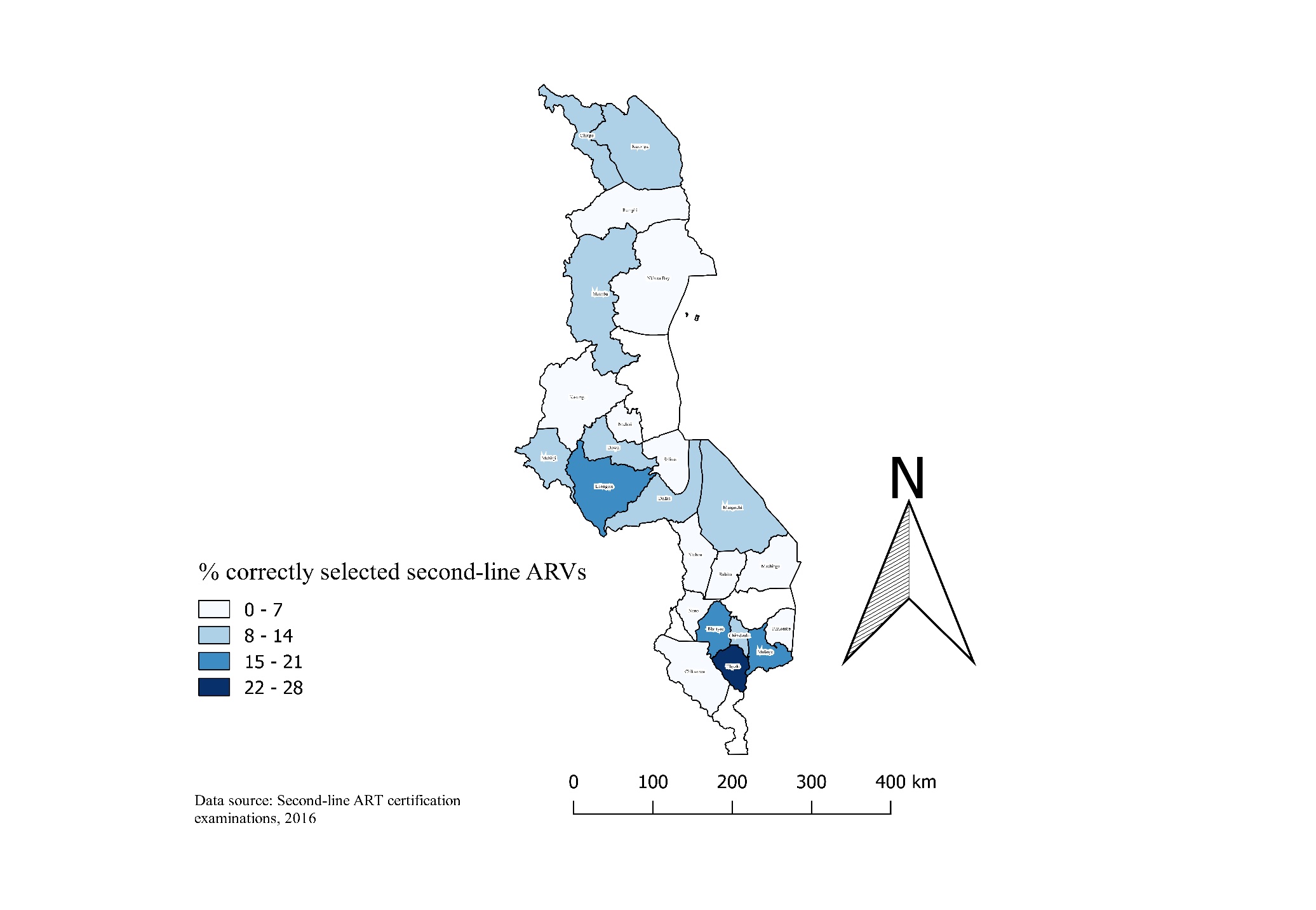

ARVs=antiretroviral drugs. We excluded the following districts due to small numbers: Zomba (n=7), Nsanje (n=3), Nkota-kota (n=1)

**Figure 4: Percentage of ART providers that correctly dosed second-line ARV regimens in Malawi in 2016 (n=743)**

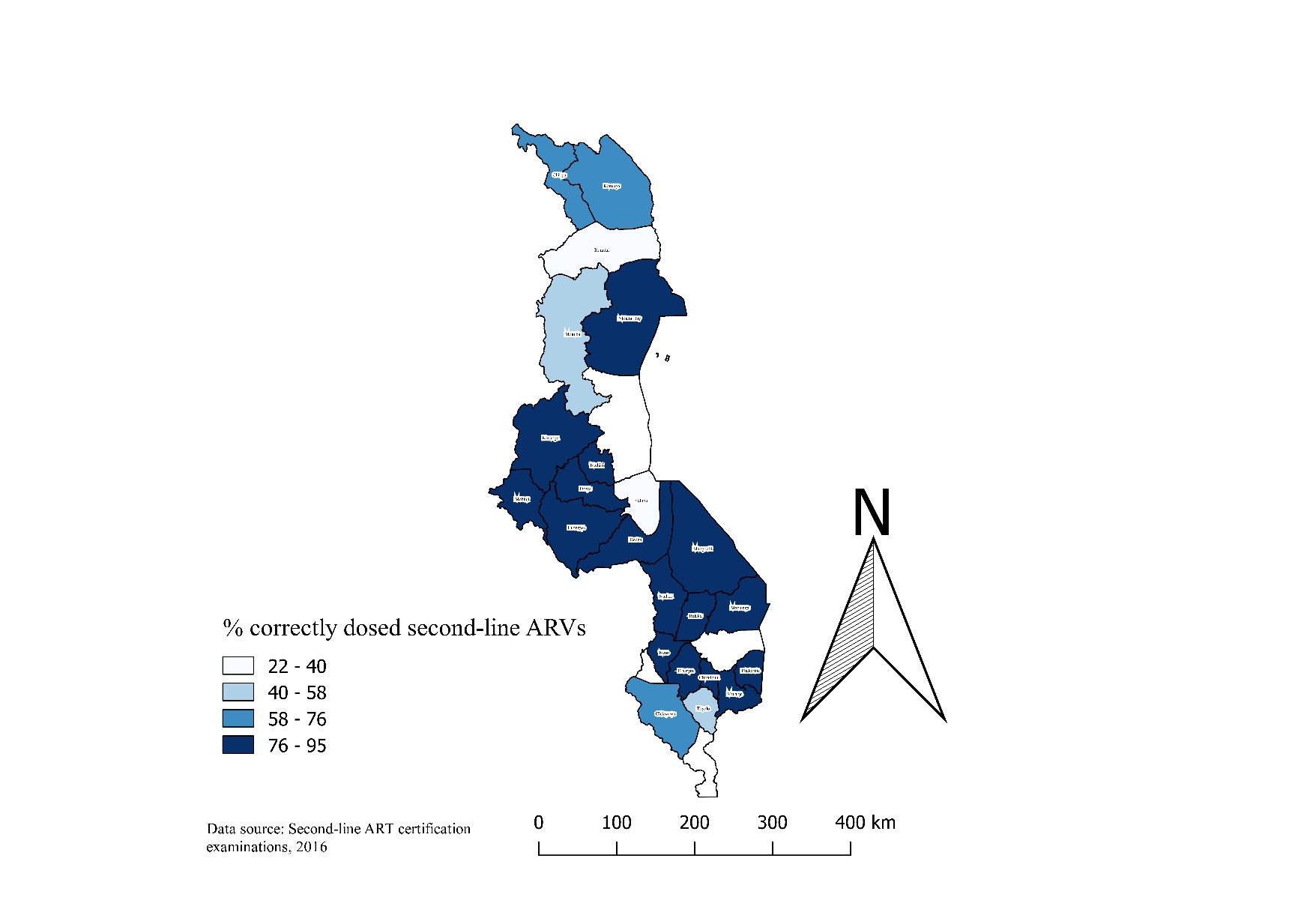

ARVs=antiretroviral drugs. We excluded the following districts due to small numbers: Zomba (n=7), Nsanje (n=3), Nkota-kota (n=1)

**Figure 5: Percentage of ART providers that understood viral load collection, interpreted viral load results and used viral load results to manage patients in Malawi in 2016 (n=743)**

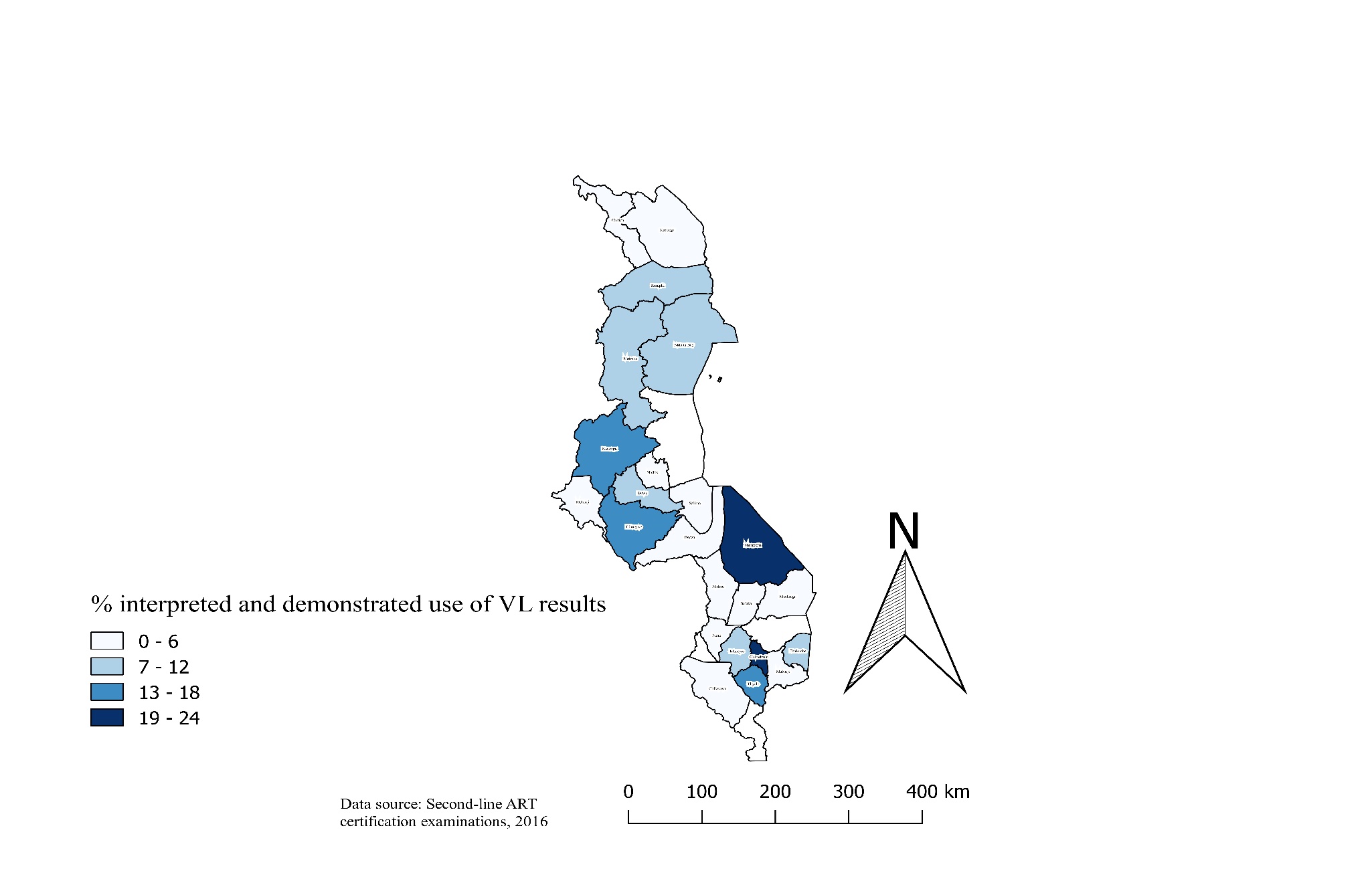

ART=antiretroviral therapy; VL=Viral load. NB: We excluded the following districts due to small numbers: Zomba (n=7), Nsanje (n=3), Nkota-kota (n=1)

**Table 1: Association of participant characteristics with percentage that passed second-line ART certification examinations in Malawi 2016**

| **Patient characteristics** | **n (%)** | **No. passed*** | **% passed (95%CI)** | **P-value**** |
| --- | --- | --- | --- | --- |
| **Total** | 754 (100) | 290 | 38.46 (34.97-42.04) |  |
| **Region** |  |  |  | 0.50 |
| *North* | 134 (18) | 46 | 34.33 (26.35-43.02) |  |
| *Central* | 220 (29) | 84 | 38.18 (31.73-44.95) |  |
| *Southern* | 400 (53) | 160 | 40.00 (35.16-44.99) |  |
| **Cadre** |  |  |  | <0.001 |
| *Medical Doctor (with at least MBBS)* | 21 (3) | 14 | 66.67 (43.03-85.41) |  |
| *Clinical Officer (with Diploma in*  *Clinical Medicine)* | 241 (32) | 136 | 56.43 (49.92 -62.79) |  |
| *Medical Assistant* | 147 (20) | 36 | 24.49 (17.78-32.26) |  |
| *Registered Nurse* | 44 (6) | 17 | 36.64 (24.36-54.50) |  |
| *Nurse Midwife Technician* | 260 (34) | 73 | 28.08 (22.70-33.96) |  |
| *Community Nurses* | 41 (5) | 14 | 34.15 (20.08-50.59) |  |

*Passing was based on getting at least 80% of the assessment examination

**Chi-Square p-values

MBBS=Bachelor of Medicine and Bachelor of Surgery

**Table 2: Association of participant characteristics with percentage that correctly answered questions in the four categories of second-line ART certification examinations in Malawi 2016**

| **Patient characteristics** | **n (%)** | **Categories of the second-line ART certification questions*** | | | | | | | | | | | |
| --- | --- | --- | --- | --- | --- | --- | --- | --- | --- | --- | --- | --- | --- |
|  |  | **Correctly identified second-line ARV regimens** | | | **Correctly selected second-line ARV regimens for managing different patients** | | | **Correctly dosed different second-line ARV regimens** | | | **Correctly interpreted viral load results** | | |
|  |  | **No. passed** | **% passed (95%CI)** | **P-value**** | **No. passed** | **% passed (95%CI)** | **P-value**** | **No. passed** | **% passed (95%CI)** | **P-value**** | **No. passed** | **% passed (95%CI)** | **P-value**** |
| **Total** | 754 (100) | 518 | 68.70 (65.26-72.00) |  | 71 | 9.42 (7.43-11.73) |  | 588 | 77.98 (74.85-80.89) |  | 67 | 8.89 (6.95-11.14) |  |
| **Region** |  |  |  | 0.06 |  |  | 0.69 |  |  | <0.001 |  |  | 0.81 |
| *North* | 134 (18) | 87 | 64.93 (56.21-72.96) |  | 10 | 7.46 (3.64-13.30) |  | 87 | 64.93 (56.21-72.96) |  | 10 | 14.29 (3.64-13.30) |  |
| *Central* | 220 (29) | 141 | 64.09 (57.37-70.43) |  | 22 | 10.00 (6.37-14.75) |  | 178 | 80.91 (75.08-85.88) |  | 20 | 7.73 (4.29-12.64) |  |
| *Southern* | 400 (53) | 290 | 72.50 (67.84-76.82) |  | 39 | 9.75 (7.03-13.09) |  | 323 | 80.75 (76.54-84.50) |  | 37 | 7.34 (4.85-10.58) |  |
| **Cadre** |  |  |  | <0.001 |  |  | <0.001 |  |  | 0.27 |  |  | <0.001 |
| *Medical Doctor (with at least MBBS)* | 21 (3) | 18 | 85.71 (63.66-96.95) |  | 7 | 33.33 (14.59-56.97) |  | 18 | 85.71 (63.66-96.95) |  | 3 | 14.29 (3.05-36.34) |  |
| *Clinical Officer (with Diploma in*  *Clinical Medicine)* | 241 (32) | 194 | 80.50 (74.92-85.30) |  | 40 | 16.60 (12.13-21.91) |  | 195 | 80.91 (75.37-85.67) |  | 36 | 14.94 (10.69-20.08) |  |
| *Medical Assistant* | 147 (20) | 91 | 61.90 (53.54-69.78) |  | 7 | 4.76 (1.94-9.57) |  | 113 | 76.87 (69.21-83.42) |  | 5 | 3.04 (1.11-7.76) |  |
| *Registered Nurse* | 44 (6) | 32 | 72.73 (57.21-85.04) |  | 0 | 0.00 (0.00-8.00) |  | 38 | 86.36 (72.65-94.82) |  | 2 | 4.55 (0.56-15.47) |  |
| *Nurse Midwife Technician* | 260 (34) | 155 | 59.62 (53.38-65.63) |  | 13 | 5.00 (2.69-8.40) |  | 192 | 73.85 (68.06-79.08) |  | 15 | 5.77 (3.26-9.34) |  |
| *Community Nurses* | 41 (5) | 28 | 68.29 (51.91-81.92) |  | 4 | 9.76 (2.72-23.13) |  | 32 | 78.05 (62.39-89.44) |  | 6 | 14.63 (5.57-29.17) |  |

*Passing for each category was based on getting 100% of the marks within each category

**Chi-Square p-values

ARV=Antiretroviral drugs

ART=Antiretroviral therapy

MBBS=Bachelor of Medicine and Bachelor of Surgery
